# Post-acute sequelae of SARS-CoV-2 (PASC) impact quality of life at 6, 12 and 18 months post-infection

**DOI:** 10.1101/2022.08.08.22278543

**Authors:** Zoe O Demko, Tong Yu, Sarika K. Mullapudi, M. Gabriela Varela Heslin, Chamia A. Dorsey, Christine B. Payton, Jeffrey A. Tornheim, Paul W. Blair, Shruti H. Mehta, David L. Thomas, Yukari C. Manabe, Annukka A.R. Antar, the OutSMART Study Team

## Abstract

Little data exist on long COVID outcomes beyond one year. In a cohort enrolled with mild-moderate acute COVID-19, a wide range of symptoms manifest at 6, 12, and 18 months. Endorsing over 3 symptoms associates with poorer quality of life in 5 domains: physical, social, fatigue, pain, and general health.

## Introduction

Millions globally currently experience or have previously experienced one or more symptoms or sequelae of long COVID, also referred to as Post-Acute Sequelae of SARS-CoV-2 (PASC).[1] Despite the large global burden, scant data exists on PASC beyond 12 months from acute infection. Here, we prospectively characterize the longitudinal course of symptoms and quality of life among a primarily non-hospitalized cohort enrolled during acute COVID-19 and evaluate the relationship between post-acute symptoms and quality of life at 6, 12, and 18 months.

## Methods

We enrolled a convenience sample of non-hospitalized adults within 48 hours of a positive SARS-CoV-2 PCR test from a large academic health system beginning in April 2020.[2] Non-hospitalized household contacts were also enrolled. Participants completed surveys one, three, six, twelve, and eighteen months post-enrollment. By May 2022, 144 post-COVID adult participants were 12+ months from acute infection, with a median 699 days since infection (IQR 665-728). We approximated infection date using symptom onset date except for one acutely asymptomatic participant, for whom first positive test date was used. 70 of 144 participants completed a survey at 12 and/or 18 months and are included here. Participants were included regardless of PASC status. Demographic characteristics of those who did and did not complete surveys are compared in Supplemental Table 1. Five participants received at least one dose of an mRNA COVID-19 vaccine prior to infection with a median 31 days since first dose (range: 2-89). Informed consent was obtained from all participants and the protocol received IRB approval.

Participants indicated the presence and severity (on a scale of 1-5) in the past week of each of 49 symptoms: 38 from the FLU-PRO© Plus and 11 additional symptoms derived from patient-led research on long COVID[3]. Participants reporting fatigue completed the Fatigue Severity Scale (FSS), and those endorsing sleep disturbances completed the Insomnia Severity Index (ISI). Mental health and quality of life (QoL) were assessed using the Generalized Anxiety Disorder 7-item (GAD-7), Personal Health Questionnaire Depression 8-item (PHQ-8), General Practitioner Assessment of Cognition (GPCOG), EuroQuol EQ-5D-5L overall health question, and 36-Item Short Form Survey (SF-36). We used the RAND 36-Item Health Survey scoring method to characterize eight domains of QoL from the SF-36 instrument (Supplemental Methods, Supplemental Table 2).

Analyses were performed using Stata 16.0[4]. T-tests or rank-sum tests were used to compare continuous variables between groups, and Pearson’s chi-squared or Fisher’s exact tests were used to compare categorical variables.

## Results

The median age of the cohort at enrollment was 53 (IQR 43-61), 61% were women, and the median BMI was 30 (IQR 24-35). 51% identified as non-Hispanic White, 23% as non-Hispanic Black or African American, and 19% as Hispanic of any race (Supplemental Table 1). At 12 months post-COVID, 67% (40 of 60) had returned to their usual pre-COVID health and 80% (48 of 60) had returned to their usual pre-COVID activities. At 18 months, 77% (27 of 35) had returned to their usual health and 83% (29 of 35) to their usual activities. Having returned to pre-COVID health was significantly associated with increased quality of life in several domains at 6 and 12 months post-COVID-19 (Table 1).

**Table 1.** Correlation between return to usual health/activities and either quality-of-life domains or mental health

|  | Month 6 (N=37) |  |  |  | <sup>a</sup> Month 12 (N=58) |  |  |  | <sup>a</sup> Month 18 (N=33) |  |  |  |
| --- | --- | --- | --- | --- | --- | --- | --- | --- | --- | --- | --- | --- |
|  | Returned Health<br>(n=30) |  | Returned Activities<br>(n=32) |  | Returned Health<br>(n=40) |  | Returned Activities<br>(n=48) |  | Returned Health<br>(n=27) |  | Returned Activities<br>(n=29) |  |
|  | Diff. in<br>Means | p-value | Diff. in<br>Means | p-value | Diff. in<br>Means | p-value | Diff. in<br>Means | p-value | Diff. in<br>Means | p-value | Diff. in<br>Means | p-value |
| **PF** | 15.59 | 0.006** | 11.81 | 0.132 | 25.40 | <0.001*** | 30.89 | <0.001*** | 0.46 | 0.632 | 18.92 | 0.161 |
| **EF** | 27.13 | 0.005** | 14.86 | 0.344 | 23.49 | <0.001*** | 27.09 | <0.001*** | -0.20 | 0.759 | 5.56 | 0.766 |
| **EW** | 10.04 | 0.073 | 0.56 | 0.857 | 8.65 | 0.053 | 16.54 | 0.004** | 2.83 | 0.366 | -3.89 | 0.888 |
| **SF** | 11.81 | 0.127 | 12.31 | 0.333 | 13.65 | 0.016* | 23.87 | 0.007** | -6.94 | 0.267 | 12.50 | 0.575 |
| **PA** | 24.62 | 0.0151* | 18.70 | 0.449 | 7.29 | 0.165 | 9.65 | 0.132 | -10.65 | 0.214 | 4.78 | 0.919 |
| **GH** | 25.40 | 0.027* | 27.78 | 0.0441* | 18.71 | 0.004** | 24.20 | 0.004** | 4.72 | 0.741 | 7.24 | 0.605 |
| **EQ-5D-5L** | 13.02 | 0.074 | 10.72 | 0.721 | 17.64 | <0.001*** | 24.09 | 0.001** | 5.38 | 0.271 | 13.31 | 0.706 |
| **GAD-7** | — | — | — | — | -0.80 | 0.573 | -2.22 | 0.279 | -1.59 | 0.485 | 1.57 | 0.652 |
| **PHQ-8** | — | — | — | — | -2.84 | 0.022* | -4.69 | 0.003** | -0.77 | 0.532 | -1.86 | 0.786 |
| **GPCOG** | 0.75 | 0.103 | 0.61 | 0.395 | 0.50 | 0.064 | 1.27 | 0.003** | -0.13 | 0.842 | 0.07 | 0.878 |

<sup>a</sup> 2 participants at 12 and 18 months were missing quality of life data and were excluded.
Diff. in Means = Difference between mean scores among those who had and those who had not returned to usual health or activities
SF-36 Quality of Life Domains: PF = Physical Functioning, EF = Energy/Fatigue, EW = Emotional Wellbeing, SF = Social
Functioning, PA = Pain, GH = General Health; EQ-5D-5L = EuroQol Quality of Life scale (general health question only); GAD-7 =
Generalized Anxiety Disorder 7-Item; PHQ-8 = Personal Health Questionnaire 8-item Depression Scale; GPCOG = General
Practitioner Assessment of Cognition (self-report section only). P-values were obtained using rank-sum tests.
\*indicates p<0.05, \*\*indicates p<0.01, \*\*\*indicates p<0.001; —indicates insufficient data.

Fatigue was the most frequently reported symptom with 38% (14 of 37), 29% (17 of 59), and 31% (11 of 36) of participants reporting fatigue at 6, 12, and 18 months post-COVID-19 (Figure 1a, Supplemental Table 3). Of the 34 participants reporting fatigue at any timepoint, 7 reported fatigue at more than one timepoint and 2 at all three timepoints, exemplifying the potential waxing/waning character of PASC symptoms. Among participants reporting fatigue, 21%, 35%, and 33% reported severe fatigue (4-5 on a scale of 1-5), at 6, 12, and 18 months, respectively. The mean (SD) Fatigue Severity Scale-9 score for participants reporting any fatigue at 6, 12, and 18 months was 3.61 (±1.68), 4.21 (±1.44), and 4.03 (±1.25) (Supplemental Table 4). This compares to mean (SD) FSS-9 scores of 3.00 (±1.08) in healthy people, 4.66 (±1.64) in people with multiple sclerosis, and 4.34 (±1.64) in people diagnosed with sleep-wake disorders in another cohort[5]. Post-exertional fatigue was present in 38% (5 of 13), 50% (8 of 16), and 17% (2 of 12) of participants reporting any fatigue at 6, 12, and 18 months post-COVID-19. 8 of the 34 participants (24%) reporting fatigue at any timepoint had pre-COVID diagnoses of depression or bipolar disorder, compared to 20% among all included participants.

**Figure 1.**
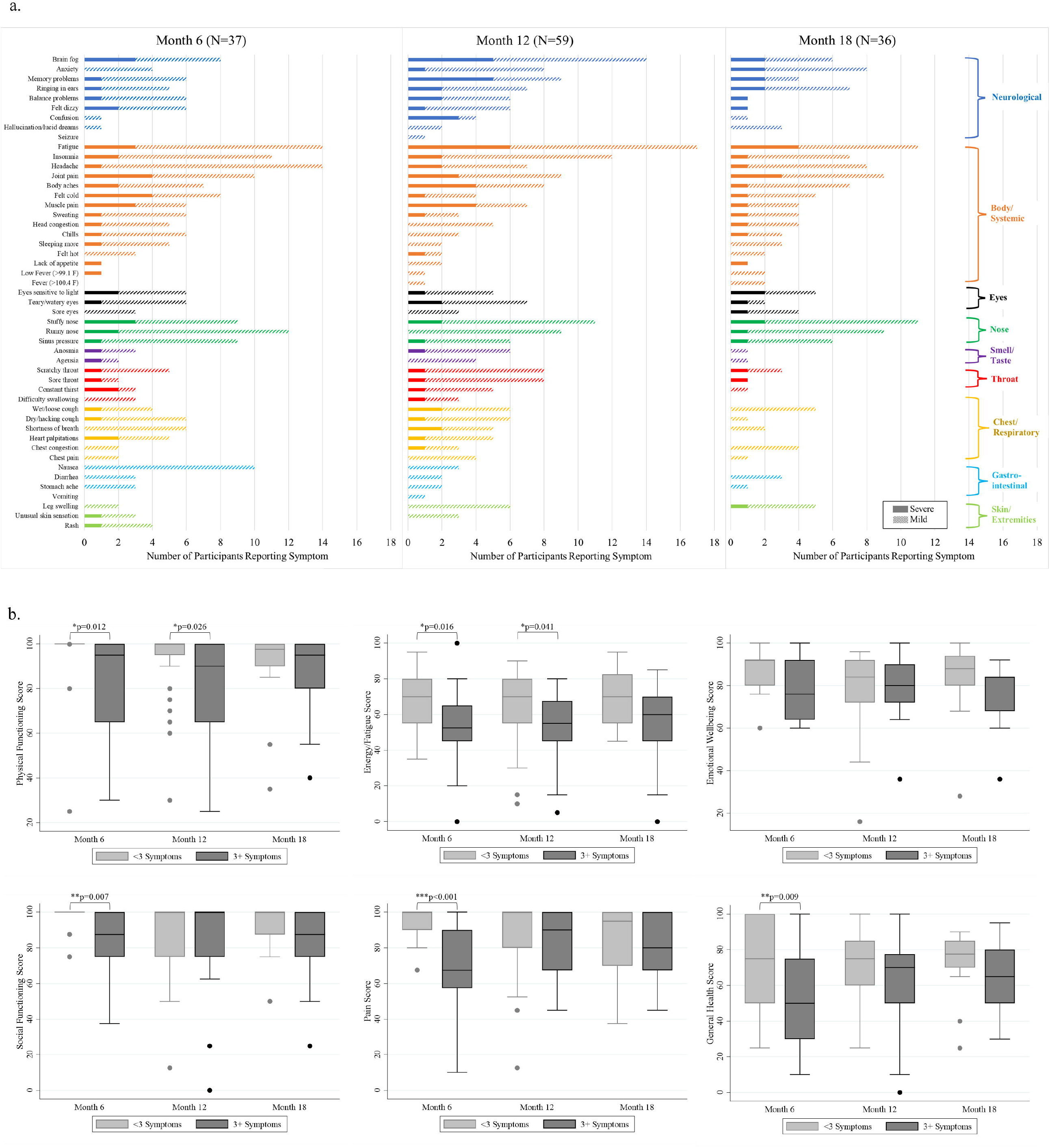
Long COVID symptoms and their association with quality of life domains a. Number of participants reporting symptoms over time, grouped by symptom domain b. SF-36 quality-of-life domain scores by number of symptoms (3+ symptoms versus <3) reported over time. Statistical significance was determined using a rank-sum test.

Insomnia and sleeping more than usual were frequently reported by participants, with 38% (14 of 37), 22% (13 of 59), and 31% (11 of 36) reporting one or both at 6, 12, and 18 months. ISI scores >10 are a valid screen for insomnia[6]. Median (IQR) ISI scores at 6, 12, and 18 months were 17 (13-20), 16 (12.5-20.5), and 14 (12-19.5), in those reporting any sleep problems, suggesting possible modest decline in insomnia severity over time post-infection. 4 of 30 participants (13%) reporting insomnia at any timepoint had baseline diagnoses of anxiety, the same proportion as that among all participants included, and 13% had prior diagnoses of depression or bipolar disorder.

Cognitive impairment was also highly reported, with 22% (8 of 37), 24% (14 of 59), and 17% (6 of 36) reporting brain fog or concentration difficulties at 6, 12, and 18 months post-infection. All participants answered 6 questions modified from the self-report section of the GPCOG indicating difficulties with memory, word-finding, or managing finances, medication, or transportation compared to just before their COVID-19 diagnosis. 30% (11 of 37), 54% (32 of 59), and 41% (14 of 34) of participants reported new, post-COVID-19 difficulties in one or more of these tasks at 6, 12, and 18 months post-COVID-19.

As 20% of participants reported diagnoses of depression or bipolar disorder and 13% reported a diagnosed anxiety disorder prior to COVID-19 diagnosis and enrollment, we evaluated PHQ-8 and GAD-7 scores stratified by these factors (Supplemental Table 5). Of participants without pre-existing depression or bipolar disorder, 18% (7 of 39) and 38% (9 of 24) had symptoms of mild depression (PHQ-8 score 5-9) at 12 and 18 months post-COVID-19, while 7.7% (3 of 39) and 0% (of 24) had symptoms of moderate to moderately-severe depression (PHQ-8 score 10-19). 2.3% (1 of 44) and 7.1% (2 of 28) of participants without pre-existing anxiety disorder had symptoms of mild anxiety (GAD-7 score 5-9), while 9.1% (4 of 44) and 3.6% (1 of 28) had symptoms of moderate to severe anxiety (GAD-7 score 10-21) at 12 and 18 months post-COVID-19.

Participants at 6, 12, and 18 months post-infection generally reported high quality of life (SF-36) scores in the physical and emotional limitation domains, indicating minimal limitation, while scores in the domains of physical functioning, energy/fatigue, emotional wellbeing, pain, and general health were lower (Figure 1b, Supplemental Table 4). Scores across the eight SF-36 QoL domains did not vary significantly between timepoints. Median (IQR) scores on the EQ-5D-5L general health question were also similar at 6, 12, and 18 months post-COVID-19: 85 (80-94), 85 (71-90), and 84 (75-95).

Participants reported that symptoms directly interfered with their daily activities: 35% (11 of 31), 22% (13 of 58), and 13% (4 of 31) at 6, 12, and 18 months post-COVID-19. Participants reporting 3+ symptoms, compared to those reporting <3 symptoms, had significantly lower scores in 5 of 8 SF-36 QoL domains (physical functioning, social functioning, energy/fatigue, pain, general health) after 6 months (Figure 1b). Physical functioning and energy/fatigue scores were significantly lower among those reporting 3+ symptoms after 12 months. There were also statistically significant associations between most QoL domains and symptom domains, particularly neurological and systemic (Supplemental Figure).

## Discussion

We demonstrate in a prospective observational cohort that a high proportion of individuals with pre-delta variant, non-severe acute COVID-19 continue to report a wide range of symptoms at 6, 12, and 18 months post-infection, and that having multiple symptoms is associated with lower quality of life over a year after infection. This data extend to 18 months previously reported PASC symptoms and associations with QoL measures at earlier time points.[7-11] Fatigue, brain fog, nasal congestion and discharge, headache, sleep disturbances, and body aches were the most common symptoms. Neurological, mood, and systemic symptoms persisted in over half of participants surveyed at 18 months post-COVID-19. Almost one-quarter of participants reported not having returned to their usual pre-COVID health by 18 months, while some reported returning to work and other activities before feeling entirely recovered from COVID-19. Symptomatic participants reported that symptoms interfered with activities up to 18 months post-infection, and several quality of life domains were statistically significantly associated with multi-symptom morbidity.

Limitations of this study include the relatively small size of the cohort, the proportion of participants who missed survey timepoints, and attrition over the follow-up period, which may lead to sampling and attrition biases. Strengths of the study include prospective enrollment during acute COVID-19, prolonged follow-up with validated survey instruments, and a primarily non-hospitalized cohort.

More research is urgently needed to characterize and address the lasting morbidity associated with long COVID, especially in the face of omicron vaccine-breakthrough cases.

## Data Availability

All data produced in the present study are available upon reasonable request to the authors.

## Acknowledgements

We would like to acknowledge Sidney Saint-Hilaire, Zihan Yang, Justin Chan, Mira Prabhu, and Tanique Bennett from the OutSMART Study Team; the CCPSEI Study Team; and the Johns Hopkins Institute for Clinical and Translational Research for their support of this work.

