## Supplemental Material for "Post-acute sequelae of SARS-CoV-2 (PASC) impact quality of life at 6, 12 and 18 months post-infection"

*Methods – Recruitment and Enrollment*

Participants were initially approached for enrollment by convenience sampling beginning in April 2020 (Phase 1), and randomized sampling of potential participants from all SARS-CoV-2 PCR positive ambulatory patients in the health system began in December 2020 (Phase 2). Concurrent recruitment of enrollees’ household contacts, including children, began in Phase 2. Both SARS-CoV-2-positive and negative contacts were eligible. Participants originally enrolled in Phase 1 were invited to re-enroll in Phase 2 if they had participated in study events by providing samples during Phase 1. Of 183 participants enrolled in Phase 1, 110 were eligible to re-enroll in Phase 2 and 72 re-enrolled. During Phase 2, 297 eligible patients were successfully contacted of whom 103 enrolled. 40 household contacts of those 103 were also enrolled for a total cohort size of 215. Informed consent was obtained from adult participants and parent/guardian consent was obtained for pediatric participants. Informed assent was obtained from pediatric participants aged 7-17 years.

As the instruments used in this analysis were validated only for the adult population, we excluded the 18 household contacts under 18 years of age, leaving 197 adult participants. 144 adult participants had reached the 12-month post-infection timepoint by May 9^th^ 2022, and 72 had reached the 18-month timepoint. 70 of the 144 who had reached 12 months completed a survey at the 12-month and/or 18-month timepoint and are included here. Of those, 37 completed surveys at 6 months, 62 completed surveys at 12 months, and 37 completed surveys at 18 months.

*Methods - Scales and Instruments:*

Survey data were collected and stored using the REDCap platform[1].

The FLU-PRO© Plus scale measures symptom severity on a 5-point Likert scale, with values including: 1 – not at all, 2 – a little bit, 3 – somewhat, 4 – quite a bit, and 5 – very much. It contains 36 symptoms consistent with influenza-like illnesses and two questions relating to loss of taste and smell relating specifically to COVID-19[2, 3]. We considered scores of 2 and 3 to indicate mild symptom severity and scores of 4 and 5 as severe.

The Fatigue Severity Score (FSS) is a 9-item questionnaire assessing patients’ perception of the impact of fatigue on their quality of life and activities of daily living, with one additional question on participants’ global fatigue level from 1 to 10 (10 indicating normal energy). Each question is scored from 1 to 7: 1 indicates strong disagreement with the statement and 7 indicates strong agreement. Total score may be calculated by summation or averaging of the nine question scores[4, 5]. We evaluated fatigue using the FSS-9 average scoring method, resulting in a score range of 7 to 63 with higher scores indicating more severe fatigue.

The Insomnia Severity Index (ISI) is a 7-item questionnaire assessing patients’ perceived insomnia symptoms and resulting disruption to their lives, as well as their degree of concern over these symptoms. Each item is scored from 1 to 5, with higher scores indicating worse symptom severity or consequences. Composite scores are calculated by summation of all questions and range from 5 to 35, with higher scores indicating worse insomnia. A score of 10 or above performs well as a screen for insomnia[6-8].

The PHQ-8 is an 8-item questionnaire measuring the frequency of symptoms of depression over the prior two weeks. Each item is scored from 0 to 3: 0 – not at all, 1 – several days, 2 – more than half the days, 3 – nearly every day. Total scores range from 0 to 24. Scores under 5 indicate no significant depressive symptoms, while 5-9 indicates mild depressive symptoms, 10-14 moderate depressive symptoms, 15-19 moderately-severe depressive symptoms, and 20-24 severe depressive symptoms[9, 10]. The GAD-7 measures anxiety symptoms using a 7-item questionnaire with answer choices identical to those on the PHQ-8, and is scored similarly with total scores ranging from 0 to 21[11]. Both the PHQ-8 and GAD-7 also include a final question asking participants to what degree their symptoms made activities of daily living difficult for them, with options including not at all, somewhat, very, and extremely[9-11].

The self-report questionnaire section of the GPCOG consists of six yes/no items relating to participants’ perceived difficulty with certain activities due to cognitive limitations as compared to a prior timepoint, with 1 indicating no difficulty and 0 indicating difficulty with a given task. Composite scores are calculated by summation of the 6 questions and range from 0 to 6. Scores of 3 or below indicate significant cognitive impairment[12]. We asked participants to evaluate their current ability to complete the tasks as compared to before they had COVID-19.

The SF-36 converts 36 Likert and binary questions relating to patients’ activities and perceptions of health status into scores for eight domains of QoL[13, 14]. We used the RAND 36-Item Health Survey 1.0 scoring method to derive scores for the following domains: physical functioning, physical limitation of role fulfillment, emotional limitation of role fulfillment, energy/fatigue, emotional wellbeing, social functioning, pain, and general health[15, 16]. Domain scores range from 0 to 100, with higher scores indicating better quality of life. The EQ-5D-5L general health question asks participants to provide one number from 0 to 100 on a visual analog scale representing their overall health[17]. All validated scales used are displayed in Supplemental Table 2.

*Results – Cohort Characteristics:*

Among the 70 participants included in this analysis, the median age was 53 years (IQR: 43, 61), 61% were female, 19% identified as Hispanic of any race, 23% as non-Hispanic Black/African American, 51% as non-Hispanic White, and 6% as other races (Supplemental Table 1). Compared to the 70 included participants, the 74 study participants who reached the Month-12 follow-up timepoint but did not complete a survey were similar in age (median 53) and sex (66% female). However, they had a significantly different racial/ethnic breakdown by Fisher’s exact test (8% Hispanic, 57% NH-Black, 31% NH-White, 4% Other, p<0.001).

*Results – Sleep disturbance:*

Two participants who reported fatigue at Month 6 and one at Month 12 did not complete all FSS questions and their scores were excluded. Two participants who reported sleep disturbances at Month 12 and one at Month 6 did not complete all ISI questions and their scores were excluded.

Of the 30 participants who reported sleep disturbances at any timepoint, 8 reported them at multiple timepoints. 2 endorsed sleep disturbances at months 6 and 12, as did 1 at months 12 and 18 and 5 at months 6 and 18. Within-person change on the ISI was highly variable, with an average decrease of 2.2 points between Months 6 and 18. This paired with the above general ISI trends across timepoints suggest a modest decline in insomnia severity over time post-infection.

Supplemental Table 1. Demographic characteristics of participants eligible for and included in cohort (eligible N=144)

| **Demographic Characteristic** | **Included (n=70)**  **N (%)** | **^a^ Excluded (n=74)**  **N (%)** | **p-value** |
| --- | --- | --- | --- |
| **Age (Median, IQR)** | 53 (43, 61) | 53 (37, 60) | 0.222 |
| **^b^ BMI (Median, IQR)** | 30 (24, 35) | 31 (25, 38) | 0.329 |
| **Sex (assigned at birth)** |  |  | 0.357 |
| Female | 43 (61.4%) | 49 (66.2%) |  |
| Male | 27 (38.6%) | 25 (33.8%) |  |
| **Race/Ethnicity** |  |  | 0.000 |
| Hispanic | 13 (18.6%) | 6 (8.1%) |  |
| Non-Hispanic Black/African American | 16 (22.9%) | 42 (56.8%) |  |
| Non-Hispanic Other | 4 (5.7%) | 3 (4.1%) |  |
| Non-Hispanic White | 36 (51.4%) | 23 (31.1%) |  |
| Prefer Not to Answer | 1 (1.4%) | 0 (0%) |  |
| **^c^ Healthcare Worker** |  |  | 0.439 |
| Yes | 25 (36.8%) | 24 (43.6%) |  |
| No | 43 (63.2%) | 31 (56.4%) |  |
| **Baseline Co-Morbidities** |  |  |  |
| ^b^ Overweight (BMI ≥ 25) | 47 (70.2%) | 55 (76.4%) | 0.406 |
| Cardiovascular disorder | 38 (54.3%) | 35 (47.3%) | 0.402 |
| Hypertension | 25 (35.7%) | 27 (36.5%) | 0.923 |
| Gastrointestinal disorder | 19 (27.1%) | 23 (31.1%) | 0.603 |
| Pulmonary disorder | 18 (25.7%) | 23 (31.1%) | 0.476 |
| Asthma/COPD/Emphysema | 11 (15.7%) | 17 (23.0%) | 0.271 |
| Endocrine disorder | 18 (25.7%) | 20 (27.0%) | 0.858 |
| Diabetes | 12 (17.1%) | 14 (18.9%) | 0.782 |
| Psychological/Mood Disorder | 19 (27.1%) | 18 (24.3%) | 0.699 |
| Renal disorder | 6 (8.6%) | 7 (9.5%) | 0.853 |
| Autoimmune disorder | 5 (7.1%) | 7 (9.5%) | 0.615 |
| Past or current cancer diagnosis | 7 (10.0%) | 3 (4.1%) | 0.161 |
| Immune suppressing medication | 7 (10.0%) | 2 (2.7%) | 0.071 |
| Transplant recipient | 2 (2.9%) | 2 (3.4%) | 0.885 |
| Hematological disorder | 1 (1.4%) | 2 (2.7%) | 0.593 |

^a^ Participants were excluded if no survey was completed at either month 12 or month 18; ^b^ Baseline BMI was missing for 3 included participants and 2 excluded participants; ^c^ Healthcare worker status was missing for 2 included participants and 11 excluded participants; p-values were obtained from t-tests or rank-sum tests (continuous variables), and Pearson’s chi-squared or Fisher’s exact tests (categorical variables)

Supplemental Table 2. Validated tools used to measure participant symptoms and quality-of-life

| **Tool** | **Abbreviation** | **Variable Measured** | **No. Items** | **Item Type(s)** | **Score Range** | **Citations** |
| --- | --- | --- | --- | --- | --- | --- |
| InFLUenza Patient-Reported Outcome  (+ COVID-19) | FLU-PRO+ | Symptom severity | 38 | 5-level Likert | 1-5 | 2, 3 |
| Fatigue Severity Score | FSS | Fatigue severity; impairment due to fatigue | 10 | 7-item Likert (9);  1 to 10 ordinal (1) | 1-9 (average)  or 9-63 (sum);  0 to 10 | 4, 5 |
| Insomnia Severity Index | ISI | Insomnia/sleep disturbance severity; impairment due to insomnia/sleep disturbance | 7 | 5-item Likert | 5 to 35 | 6, 7, 8 |
| Patient Health Questionnaire Depression 8-Item Scale | PHQ-8 | Depressive symptoms; impairment due to depression | 9 | 4-item Likert | 0 to 24 | 9, 10 |
| Generalized Anxiety Disorder 7-Item Scale | GAD-7 | Anxiety symptoms, impairment due to anxiety | 8 | 4-item Likert | 0 to 21 | 11 |
| General Practitioner Assessment of Cognition | GPCOG | Cognitive function | 6 | Yes/No | 0 to 6 | 12 |
| EuroQol 5-Dimension 5-Level Visual Analog Scale | EQ-5D-5L VAS | Overall health | 1 | Visual analog scale | 0 to 100 | 17 |
| 36-Item Short Form Survey | SF-36 | Quality-of-life across 8 domains | 36 | 3-level Likert (10)  5-level Likert (9)  6-level Likert (10)  Yes/No (7) | 0 to 100 in each of 8 domains | 13, 14, 15, 16 |

Supplemental Table 3 – Number of participants reporting symptoms at each timepoint, by severity and symptom domain

|  |  | **Month 6 (N=36)** | | | | **Month 12 (N=54)** | | | | **Month 18 (N=36)** | | | | |
| --- | --- | --- | --- | --- | --- | --- | --- | --- | --- | --- | --- | --- | --- | --- |
| **Symptom Domain** | **Symptom** | **None** | **Mild** | **Severe** | **Mild + Severe** | **None** | **Mild** | **Severe** | **Mild + Severe** | | **None** | **Mild** | **Severe** | **Mild + Severe** |
| Neurological | Brain fog | 29 | 5 | 3 | 8 | 45 | 9 | 5 | 14 | | 36 | 0 | 0 | 0 |
|  | Anxiety | 33 | 4 | 0 | 4 | 51 | 7 | 1 | 8 | | 36 | 0 | 0 | 0 |
|  | Memory problems | 31 | 5 | 1 | 6 | 50 | 4 | 5 | 9 | | 31 | 4 | 1 | 5 |
|  | Ringing in ears | 32 | 4 | 1 | 5 | 52 | 5 | 2 | 7 | | 36 | 0 | 0 | 0 |
|  | Balance problems | 31 | 5 | 1 | 6 | 53 | 4 | 2 | 6 | | 35 | 1 | 0 | 1 |
|  | Felt dizzy | 31 | 4 | 2 | 6 | 53 | 5 | 1 | 6 | | 33 | 3 | 0 | 3 |
|  | Confusion | 36 | 1 | 0 | 1 | 55 | 1 | 3 | 4 | | 36 | 0 | 0 | 0 |
|  | Hallucination/lucid dreams | 36 | 1 | 0 | 1 | 57 | 2 | 0 | 2 | | 35 | 1 | 0 | 1 |
|  | Seizure | 37 | 0 | 0 | 0 | 58 | 1 | 0 | 1 | | 32 | 4 | 0 | 4 |
| Body/Systemic | Fatigue | 23 | 11 | 3 | 14 | 42 | 11 | 6 | 17 | | 36 | 0 | 0 | 0 |
|  | Insomnia | 26 | 9 | 2 | 11 | 47 | 10 | 2 | 12 | | 34 | 2 | 0 | 2 |
|  | Headache | 23 | 13 | 1 | 14 | 52 | 5 | 2 | 7 | | 35 | 1 | 0 | 1 |
|  | Joint pain | 27 | 6 | 4 | 10 | 50 | 6 | 3 | 9 | | 31 | 5 | 0 | 5 |
|  | Body aches | 30 | 5 | 2 | 7 | 51 | 4 | 4 | 8 | | 36 | 0 | 0 | 0 |
|  | Felt cold | 29 | 4 | 4 | 8 | 55 | 3 | 1 | 4 | | 35 | 1 | 0 | 1 |
|  | Muscle pain | 31 | 3 | 3 | 6 | 52 | 3 | 4 | 7 | | 35 | 0 | 1 | 1 |
|  | Sweating | 31 | 5 | 1 | 6 | 56 | 2 | 1 | 3 | | 33 | 2 | 1 | 3 |
|  | Head congestion | 32 | 4 | 1 | 5 | 54 | 5 | 0 | 5 | | 35 | 1 | 0 | 1 |
|  | Chills | 31 | 5 | 1 | 6 | 56 | 3 | 0 | 3 | | 35 | 1 | 0 | 1 |
|  | Sleeping more | 32 | 4 | 1 | 5 | 57 | 2 | 0 | 2 | | 30 | 5 | 1 | 6 |
|  | Felt hot | 34 | 3 | 0 | 3 | 57 | 1 | 1 | 2 | | 27 | 8 | 1 | 9 |
|  | Lack of appetite | 36 | 0 | 1 | 1 | 57 | 2 | 0 | 2 | | 25 | 9 | 2 | 11 |
|  | Low Fever (>99.1 F) | 36 | 0 | 1 | 1 | 59 | 1 | 0 | 1 | | 32 | 3 | 1 | 4 |
|  | Fever (>100.4 F) | 37 | 0 | 0 | 0 | 59 | 1 | 0 | 1 | | 34 | 1 | 1 | 2 |
| Eyes | Eyes sensitive to light | 31 | 4 | 2 | 6 | 54 | 4 | 1 | 5 | | 31 | 3 | 2 | 5 |
|  | Teary/watery eyes | 31 | 5 | 1 | 6 | 52 | 5 | 2 | 7 | | 34 | 2 | 0 | 2 |
|  | Sore eyes | 34 | 3 | 0 | 3 | 56 | 3 | 0 | 3 | | 34 | 2 | 0 | 2 |
| Nose | Stuffy nose | 28 | 6 | 3 | 9 | 48 | 9 | 2 | 11 | | 35 | 0 | 1 | 1 |
|  | Runny nose | 25 | 10 | 2 | 12 | 50 | 9 | 0 | 9 | | 34 | 2 | 0 | 2 |
|  | Sinus pressure | 28 | 8 | 1 | 9 | 53 | 5 | 1 | 6 | | 33 | 3 | 0 | 3 |
| Smell/ Taste | Anosmia | 34 | 2 | 1 | 3 | 53 | 5 | 1 | 6 | | 33 | 2 | 1 | 3 |
|  | Ageusia | 35 | 1 | 1 | 2 | 55 | 4 | 0 | 4 | | 32 | 3 | 1 | 4 |
| Throat | Scratchy throat | 32 | 4 | 1 | 5 | 51 | 7 | 1 | 8 | | 31 | 3 | 1 | 4 |
|  | Sore throat | 35 | 1 | 1 | 2 | 51 | 7 | 1 | 8 | | 32 | 3 | 1 | 4 |
|  | Constant thirst | 34 | 1 | 2 | 3 | 54 | 4 | 1 | 5 | | 31 | 4 | 1 | 5 |
|  | Difficulty swallowing | 34 | 3 | 0 | 3 | 56 | 2 | 1 | 3 | | 29 | 6 | 1 | 7 |
| Chest/Respiratory | Wet/loose cough | 33 | 3 | 1 | 4 | 53 | 4 | 2 | 6 | | 27 | 6 | 3 | 9 |
|  | Dry/hacking cough | 31 | 5 | 1 | 6 | 53 | 5 | 1 | 6 | | 28 | 7 | 1 | 8 |
|  | Shortness of breath | 31 | 6 | 0 | 6 | 54 | 3 | 2 | 5 | | 29 | 6 | 1 | 7 |
|  | Heart palpitations | 32 | 3 | 2 | 5 | 54 | 4 | 1 | 5 | | 25 | 7 | 4 | 11 |
|  | Chest congestion | 35 | 2 | 0 | 2 | 56 | 2 | 1 | 3 | | 36 | 0 | 0 | 0 |
|  | Chest pain | 35 | 2 | 0 | 2 | 55 | 4 | 0 | 4 | | 33 | 3 | 0 | 3 |
| Gastro- intestinal | Nausea | 27 | 10 | 0 | 10 | 56 | 3 | 0 | 3 | | 35 | 1 | 0 | 1 |
|  | Diarrhea | 34 | 3 | 0 | 3 | 57 | 2 | 0 | 2 | | 35 | 0 | 1 | 1 |
|  | Stomach ache | 34 | 3 | 0 | 3 | 57 | 2 | 0 | 2 | | 35 | 0 | 1 | 1 |
|  | Vomiting | 37 | 0 | 0 | 0 | 58 | 1 | 0 | 1 | | 29 | 5 | 2 | 7 |
| Skin/Extre-mities | Leg swelling | 35 | 2 | 0 | 2 | 53 | 6 | 0 | 6 | | 32 | 2 | 2 | 4 |
|  | Unusual skin sensation | 34 | 2 | 1 | 3 | 56 | 3 | 0 | 3 | | 28 | 6 | 2 | 8 |
|  | Rash | 33 | 3 | 1 | 4 | 59 | 0 | 0 | 0 | | 30 | 4 | 2 | 6 |

Supplemental Table 4. Measures of central tendency for quality of life and mental health by timepoint

|  | **Month 6** | | **Month 12** | | **Month 18** | |
| --- | --- | --- | --- | --- | --- | --- |
| **Quality of Life Metric** | **N** | **Median (IQR) or n (%)** | **N** | **Median (IQR) or n (%)** | **N** | **Median (IQR) or n (%)** |
| **SF-36 Domains** |  |  |  |  |  |  |
| Physical Functioning (PF) | 37 | 100 (80, 100) | 58 | 100 (80, 100) | 34 | 95 (85, 100) |
| Physical Limitation (PL) | 37 | 100 (100, 100) | 58 | 100 (100, 100) | 34 | 100 (100, 100) |
| Emotional Limitation (EL) | 37 | 100 (100, 100) | 58 | 100 (100, 100) | 34 | 100 (100, 100) |
| Energy/Fatigue (EF) | 37 | 60 (50, 75) | 60 | 60 (50, 75) | 32 | 65 (48, 75) |
| Emotional Wellbeing (EW) | 37 | 84 (72, 92) | 60 | 84 (72, 92) | 32 | 84 (74, 92) |
| Social Functioning (SF) | 37 | 100 (87, 100) | 58 | 100 (75, 100) | 34 | 100 (75, 100) |
| Pain (PA) | 37 | 90 (68, 100) | 58 | 90 (68, 100) | 34 | 90 (70, 100) |
| General Health (GH) | 37 | 50 (50, 85) | 57 | 70 (55, 80) | 34 | 70 (50, 85) |
| **EQ-5D-5L General Health** | 37 | 85 (80, 94) | 58 | 85 (71, 90) | 30 | 84 (75, 95) |
| **GPCOG** | 37 |  | 59 |  | 34 |  |
| Composite Score |  | 6 (5, 6) |  | 5 (4, 6) |  | 6 (4, 6) |
| Score <6 |  | 11 (30%) |  | 32 (54%) |  | 14 (41%) |
| Score <4 (Impaired) |  | 4 (11%) |  | 13 (22%) |  | 4 (12%) |
| **FSS** |  |  |  |  |  |  |
| FSS-9 Score (median, IQR) | 12 | 2.9 (1.9, 3.9) | 16 | 3.6 (2.8, 5.3) | 12 | 3.9 (2.6, 4.4) |
| FSS-9 Score (mean, SD) | 12 | 3.61 (±1.68) | 16 | 4.21 (±1.44) | 12 | 4.03 (±1.25) |
| Global Fatigue Rating | 14 | 4.5 (3.3, 6) | 16 | 6 (5, 8) | 12 | 6.5 (5.8, 7) |
| **ISI Score** | 13 | 17 (13, 20) | 11 | 16 (12.5, 20.5) | 11 | 14 (12, 19.5) |

Supplemental Table 5 – GAD-7 and PHQ-8 scores stratified by prior diagnoses of anxiety, depression or bipolar disorder

|  | **Month 6** | | | | **Month 12** | | | | **Month 18** | | | |
| --- | --- | --- | --- | --- | --- | --- | --- | --- | --- | --- | --- | --- |
|  | **Anxiety** | | **No Anxiety** | | **Anxiety** | | **No Anxiety** | | **Anxiety** | | **No Anxiety** | |
| **Mental Health Evaluation Component** | **N** | **Med. (IQR) or n (%)** | **N** | **Med. (IQR) or n (%)** | **N** | **Med. (IQR) or n (%)** | **N** | **Med. (IQR) or n (%)** | **N** | **Med. (IQR) or n (%)** | **N** | **Med. (IQR) or n (%)** |
| **GAD-7** |  |  |  |  |  |  |  |  |  |  |  |  |
| Raw Score | 0 | — | 11 | 0 (0,0) | 4 | 4 (1,7) | 44 | 0 (0,2) | 5 | 5 (4,5) | 28 | 0 (0,2) |
| Score Category | 0 |  | 9 |  | 6 |  | 44 |  | 5 |  | 28 |  |
| Minimal Anxiety |  | — |  | 11 (100%) |  | 3 (50%) |  | 39 (89%) |  | 2 (40%) |  | 25 (89%) |
| Mild Anxiety |  | — |  | 0 (0%) |  | 2 (33%) |  | 1 (2%) |  | 2 (40%) |  | 2 (7%) |
| Moderate Anxiety |  | — |  | 0 (0%) |  | 1 (17%) |  | 2 (5%) |  | 1 (20%) |  | 1 (4%) |
| Severe Anxiety |  | — |  | 0 (0%) |  | 0 (0%) |  | 2 (5%) |  | 0 (0%) |  | 0 (0%) |
| Difficulty Category | 0 |  | 2 |  | 5 |  | 16 |  | 5 |  | 13 |  |
| Not Difficult at All |  | — |  | 2 (100%) |  | 2 (40%) |  | 8 (50%) |  | 1 (20%) |  | 7 (54%) |
| Somewhat Difficult |  | — |  | 0 (0%) |  | 2 (40%) |  | 6 (38%) |  | 3 (60%) |  | 6 (46%) |
| Very Difficult |  | — |  | 0 (0%) |  | 1 (20%) |  | 2 (13%) |  | 1 (20%) |  | 0 (0%) |
|  | **Depression/BPD** | | **No Depression/BPD** | | **Depression/BPD** | | **No Depression/BPD** | | **Depression/BPD** | | **No Depression/BPD** | |
| **PHQ-8** |  |  |  |  |  |  |  |  |  |  |  |  |
| Raw Score | 2 | 0 (0,0) | 9 | 1 (0,1) | 8 | 3.5 (1,6) | 39 | 1 (0,5) | 9 | 2 (1,4) | 24 | 0.5 (0,5) |
| Score Category | 2 |  | 9 |  | 8 |  | 39 |  | 9 |  | 24 |  |
| No Significant Depressive Symptoms |  | 2 (100%) |  | 8 (89%) |  | 5 (63%) |  | 29 (74%) |  | 7 (78%) |  | 15 (63%) |
| Mild Depression |  | 0 (0%) |  | 1 (11%) |  | 2 (25%) |  | 7 (18%) |  | 1 (11%) |  | 9 (38%) |
| Moderate Depression |  | 0 (0%) |  | 0 (0%) |  | 1 (13%) |  | 1 (3%) |  | 1 (11%) |  | 0 (0%) |
| Moderately Severe Depression |  | 0 (0%) |  | 0 (0%) |  | 0 (0%) |  | 2 (5%) |  | 0 (0%) |  | 0 (0%) |
| Difficulty Score | 0 |  | 4 |  | 7 |  | 21 |  | 7 |  | 12 |  |
| Not Difficult at All |  | — |  | 2 (50%) |  | 4 (57%) |  | 12 (50%) |  | 4 (57%) |  | 5 (42%) |
| Somewhat Difficult |  | — |  | 2 (50%) |  | 2 (29%) |  | 6 (32%) |  | 2 (29%) |  | 6 (50%) |
| Very Difficult |  | — |  | 0 (0%) |  | 1 (14%) |  | 3 (18%) |  | 1 (14%) |  | 1 (8%) |

*Supplemental Figure. Heat map of correlations between positive symptom domain reporting and quality of life, mental health*

|  | Month 6 (N=36) | | | | | | | | | | Month 12 (N=56) | | | | | | | | | | Month 18 (N=34) | | | | | | | | | |
| --- | --- | --- | --- | --- | --- | --- | --- | --- | --- | --- | --- | --- | --- | --- | --- | --- | --- | --- | --- | --- | --- | --- | --- | --- | --- | --- | --- | --- | --- | --- |
|  | FP | EF | EW | SF | PA | GH | EQ-5D-5L | GAD-7 | PHQ-8 | GPCOG | FP | EF | EW | SF | PA | GH | EQ-5D-5L | GAD-7 | PHQ-8 | GPCOG | FP | EF | EW | SF | PA | GH | EQ-5D-5L | GAD-7 | PHQ-8 | GPCOG |
| Neurological  (n=17, 19, 16) |  |  |  |  |  |  |  | — | — |  |  |  |  |  |  |  |  |  |  |  |  |  |  |  |  |  |  |  |  |  |
| Body/Systemic (n=24, 20, 19) |  |  |  |  |  |  |  | — | — |  |  |  |  |  |  |  |  |  |  |  |  |  |  |  |  |  |  |  |  |  |
| Eye  (n=9, 7, 8) |  |  |  |  |  |  |  | — | — |  |  |  |  |  |  |  |  |  |  |  |  |  |  |  |  |  |  |  |  |  |
| Nose  (n=18, 15, 16) |  |  |  |  |  |  |  | — | — |  |  |  |  |  |  |  |  |  |  |  |  |  |  |  |  |  |  |  |  |  |
| Throat  (n=9, 13, 4) |  |  |  |  |  |  |  | — | — |  |  |  |  |  |  |  |  |  |  |  |  |  |  |  |  |  |  |  |  |  |
| Smell/Taste  (n=3, 6, 1) |  |  |  |  |  |  |  | — | — |  |  |  |  |  |  |  |  |  |  |  | — | — | — | — | — | — | — | — | — | — |
| Chest/Respiratory (n=15, 14, 8) |  |  |  |  |  |  |  | — | — |  |  |  |  |  |  |  |  |  |  |  |  |  |  |  |  |  |  |  |  |  |
| Gastrointestinal (n=11, 4, 3) |  |  |  |  |  |  |  | — | — |  |  |  |  |  |  |  |  |  |  |  |  |  |  |  |  |  |  |  |  |  |
| Skin/Extremities (n=7, 5, 5) |  |  |  |  |  |  |  | — | — |  |  |  |  |  |  |  |  |  |  |  |  |  |  |  |  |  |  |  |  |  |

SF-36 Quality of Life Domains: PF = Physical Functioning, EF = Energy/Fatigue, EW = Emotional Wellbeing, SF = Social Functioning, PA = Pain, GH = General Health; EQ-5D-5L = EuroQol Quality of Life scale (general health question only); GAD-7 = Generalized Anxiety Disorder 7-Item; PHQ-8 = Personal Health Questionnaire Depression Scale; GPCOG = General Practitioner Assessment of Cognition. P-values were obtained using rank-sum tests.

| p≥0.1 | p<0.1 | p<0.05 | p<0.01 | p<0.001 | — indicates insufficient data |
| --- | --- | --- | --- | --- | --- |
